# Defining the Critical Components of Informed Consent for Genetic Testing: A Delphi Study

**DOI:** 10.1101/2021.06.24.21259406

**Authors:** Kelly E. Ormond, Maia Borensztein, Miranda L.G. Hallquist, Adam H. Buchanan, W. Andrew Faucett, Holly L. Peay, Maureen E. Smith, Eric P. Tricou, Wendy R. Uhlmann, Karen E. Wain, Curtis R. Coughlin, On behalf of the Clinical Genome CADRe Workgroup

## Abstract

**Purpose:** Informed consent for genetic testing has historically happened during pre-test genetic counseling, without specific guidance defining which core concepts are required.

**Methods:** The Clinical Genome Resource (ClinGen) Consent and Disclosure Recommendations Workgroup (CADRe) used a modified Expert Delphi consensus process to identify the core concepts essential to consent for clinical genetic testing. A literature review identified 77 concepts that are included in informed consent for genetic tests. Twenty five experts (9 medical geneticists, 8 genetic counselors, and 9 bioethicists) completed two rounds of Delphi surveys ranking concepts’ importance to informed consent.

**Results:** The most highly ranked concepts included: (1) genetic testing is voluntary; (2) why the test is recommended and what does it test for; (3) what results will be returned and to whom; (4) are there other types of potential results; what choices exist; (5) how will prognosis and management be impacted by results; (6) what is the potential family impact; (7) what are the test limitations and next steps; (8) potential risk for genetic discrimination and legal protections.

**Conclusion:** Defining the core concepts necessary for informed consent for genetic testing provides a foundation for quality patient care across a variety of healthcare providers and clinical indications.

## INTRODUCTION

The principle of informed consent is foundational to the premise that patients and research participants can make autonomous decisions about whether to undergo genetic testing. Critical to this process is that patients and/or research participants must receive and comprehend some specific information in order to make an informed decision and to provide consent. Marteau et al term this ‘relevant knowledge’ and define it as one of the three components of informed choice ^1^. But various standards exist in determining which information is relevant to provide to the majority of patients. The first and oldest was a “medical providers standard;” this is defined by professional guidelines that define local or national standards of what information to include as part of the informed consent process. Guidelines for Huntington Disease testing were among the first that were endorsed by providers ^2^ and advocacy organizations (http://hdsa.org/wp-content/uploads/2015/02/HDSA-Gen-Testing-Protocol-for-HD.pdf). While focused on the entire genetic counseling process, which goes beyond informed consent, some examples of professional guidelines specific to genetic testing include National Society of Genetic Counseling (NSGC) Practice Guidelines [e.g. for Alzheimer disease ^3^, *FMR1* ^4^, Neurofibromatosis ^5^], and guidelines by the American College of Obstetricians and Gynecologists (ACOG) ^6^, American Society of Clinical Oncologists (ASCO) ^7^, the American Heart Association (AHA) ^8^ or Heart Failure Society (HFS) ^9^.

In contrast, the legal case *Canterbury vs. Spence* 1972 ^10, 11^ proposed the notion of the “reasonable patient” standard. Driven by patient and family stakeholder views, this approach focuses on the amount and type of information that an average or typical patient would desire and expect to know ^12–15^. Regardless of who defines the relevant content for a typical informed consent conversation, and whether it is a clinical or research setting, there is some general consensus that patients are often overwhelmed by the large volume of educational information that is conveyed and that they frequently find it irrelevant to their questions ^16^. This is true both for patients with lower health literacy who may be overwhelmed by the information, and also, in our experience, for patients with high health literacy who may focus on information that isn’t central to the decision. In response to this, more than two decades ago, a “generic” consent model for carrier testing for reproductive risk information was proposed in order to provide sufficient information without overburdening patients ^17,18^. As genetic testing panels have expanded in recent years, additional models have been proposed to help address the ‘information overload’ issue; these include a “tiered and binned” model that was proposed in the context of multiplex cancer susceptibility testing and a similar “tiered-layered-staged” model for personal genome testing ^19–21 22^. Each of these models tries to find ways to reduce information in general, often by categorizing information either in conceptual ways (e.g. genetic test results can provide information that may vary in terms of the specific risks, including conditions beyond the differential diagnosis, and may be based on context such as personal and family history), or based on categories of potential results (e.g. medically actionable or not; low, medium or high risk).

Despite this existing literature, there remains no clear agreement either professionally or with patient stakeholders about what concepts are truly critical to allow a patient to meaningfully consent to undergo (or decline) genetic testing. The Consent and Disclosure Recommendations (CADRe) workgroup of the NIH funded Clinical Genome Resource project (clinicalgenome.org) has been focused on proposing and standardizing approaches to pre-test communication discussions beyond a traditional genetic counseling approach (e.g. targeted discussion and brief communications) and determining in which testing indications and genes/conditions they could be offered (e.g. ^23,24^. The next step in our work, through the present study, is to find consensus on these key elements (the minimum necessary and critical concepts to include in an informed consent process) to inform genetic testing practice.

## MATERIALS AND METHODS

This study used a modified Expert Delphi consensus process to define the critical concepts required for informed consent for genetic testing in a clinical setting. The Stanford University Institutional Review Board (IRB) reviewed this study and declared it exempt.

### Delphi Survey Methodology

The Delphi method, developed in the 1950’s by the Rand Corporation, is an expert consensus method that is used to systematically determine areas of agreement and disagreement around a particular topic ^25^. The method typically involves multiple phases. First, a “gathering” phase, when a list of concepts is developed, either through focus groups, a literature review, or a combination of approaches. Next, several rounds of surveys are distributed to the expert respondents in order to prioritize the concepts, provide feedback on the group’s scoring, and then allow revision of scoring to take into account the anonymous feedback from the group. Anonymity and expert status are both critical to the process. Experts bring content knowledge to the prioritization, and the anonymity aspect allows equal voice to each participant, so that the more vocal or more esteemed experts are not given more weight than other participants. Finally, analysis determines which concepts have developed consensus within the process.

### Literature Review

We completed a literature review to develop a comprehensive list of concepts involved in the consent process for genetic testing. The review identified publications listed in PubMed before June 2019 using keywords “informed consent,” “consent,” “consent form,” and “genetic [or] genomic” AND “counseling.” We then limited the papers reviewed to those that covered germline genetic testing in clinical and research settings and focused only on those that described the content of either consent forms or of genetic counseling that was part of the informed consent process. We also reviewed relevant practice guidelines, consent forms from clinical laboratories offering exome testing, and direct-to-consumer genetic testing company websites’ educational materials. The concepts were extracted from the results of the literature review, combined with concepts extracted from prior focus groups with both patients and providers that were conducted by the Clinical Genome project’s CADRe workgroup ^23^ and discussed by members of the CADRe workgroup. This resulted in a list of 77 concepts across five broad categories: (1) scope of the test (N=20), (2) description of the test and logistics (N=28), (3) confidentiality and privacy (N=13), (4) potential benefits (N=11), and (5) potential risks (N=5).

### Recruitment

A critical feature of the Delphi process is clearly defining the expert participant population, in this case, individuals who were considered experts in the topic of informed consent for genetic testing. In this study, experts were defined as genetics clinicians (genetic counselors and medical geneticists) or bioethicists with at least 5 years of experience in obtaining informed consent and results disclosure of genetic testing results, or research experience in the ethical, legal, and social implications (ELSI) of genetic testing who practiced in the United States. Experts were identified through their membership in one of three areas: (1) ethics related committees or (2) leadership participation of various clinical special interest groups within genetics organizations including the National Society of Genetic Counselors (NSGC), American College of Medical Genetics and Genomics (ACMG), American Society of Human Genetics (ASHG), and Clinical Genome Resource (ClinGen), or (3) genetics special interest groups within non-genetics organizations (e.g., American Heart Association and the American Society of Clinical Oncology (ASCO). Additional ELSI thought leaders in this area were identified by those who authored multiple publications identified in the literature review. Delphi participants were provided $75 per each completed round of surveys, for a maximum of $150 each.

### Methodology

Between November 2019 and February 2020, we conducted two rounds of Delphi surveys using Qualtrics online survey administration software; surveys were linked using a codename created by each participant at the end of Survey 1. We initially selected a two-round approach based on data that suggests two-three rounds of surveys are adequate to obtain consensus ^25^.

In Survey 1 (Supplemental Materials 1), participants were asked to rate each of the 77 concepts on a 5-point Likert scale with regards to how critical the topic is in obtaining informed consent to undertake a genetic test in a clinical setting. After ranking every item within a category, they then ranked the top five most important items within the category. This was selected as a way to differentiate preferences in a scenario where a significant number of concepts received the highest ratings. After completing ratings, rankings, and open-ended questions for all five categories, participants were asked to provide additional comments about the survey as a whole, including identifying any concepts they felt were missing from the original concept list.

Before completing Survey 2, approximately 2 months later, participants were given their individual results from Survey 1 and a summary of the group results. Participants were asked to re-rate all 77 concepts in light of reviewing this data, and rated an additional 12 concepts that were suggested by participants during Survey 1. Participants were asked to comment on a list of the 10 most highly rated concepts and whether they thought the concepts included “the minimum necessary and critical information to aid an individual in making an informed decision/consent,” and, if not, which concepts they felt were missing or could be removed.

They were subsequently shown a list of 18 ‘second tier’ concepts and asked if they felt any of these issues warranted inclusion on a core concept list and why. Lastly, participants piloted and provided open ended comments on four consent scenarios that were intended to be used for a national survey following up the Expert Delphi Surveys (Borensztein et al., 2020). Comments from these scenarios were used to inform discussions as the workgroup developed the ‘final concept list’ in this study.

### Delphi Analysis

In order to define which concepts were most highly rated by the expert participants on Survey 1, we used SPSS version 26.0 (2019) to calculate means, standard deviations and modes for each concept. In Survey 1, we also tallied the number of times a concept was ranked in the top five within a category. For the “potential risks” category, since there were only five items, we tracked the top three items. Open ended comments from both surveys were reviewed and discussed by the entire CADRe workgroup in order to determine which concepts were most highly endorsed to include in the subsequent survey.

We created a list of the top concepts from Survey 1 based on items that scored a mean >4.0 (out of 5.0) and were also frequently ranked in the top five lists. To ensure that we did not miss any critical concepts with these cutoffs, we created a second tier of concepts that met one but not both of the criteria above; for example, concepts that either scored between 3.75-3.99 but were frequently ranked in the top five, or had a score >4.0 but were not frequently ranked in the top five.

To identify the consensus list of core concepts that would allow “the minimum necessary and critical information to aid an individual in making an informed decision/consent,” we examined the mean responses for both surveys, using a combination of the criteria described above and looking for consistency across both surveys, the open ended comments from participants, and discussions within the CADRe workgroup. Some items were combined to avoid repetition. Table 2 provides examples of the manner in which this was done.

## RESULTS

One-hundred and thirty-two potentially eligible experts were identified. After excluding 36 individuals who were members of the ClinGen Steering Committee or who were genetics laboratory directors without patient-facing roles, and eight without publically available emails, 88 potential participants were emailed in November 2019. Three emails were undeliverable, and 32 confirmed their interest through an eligibility survey (32/85, or 37.6% initial response rate), one of whom was ineligible based on years of experience. Ultimately, 25 individuals (nine medical geneticists, eight genetic counselors, and nine bioethicists) completed Survey 1 (Table 1; 80% of those who expressed initial interest and were eligible), and 23 completed Survey 2, representing a 92% retention rate.

**Table 1:**
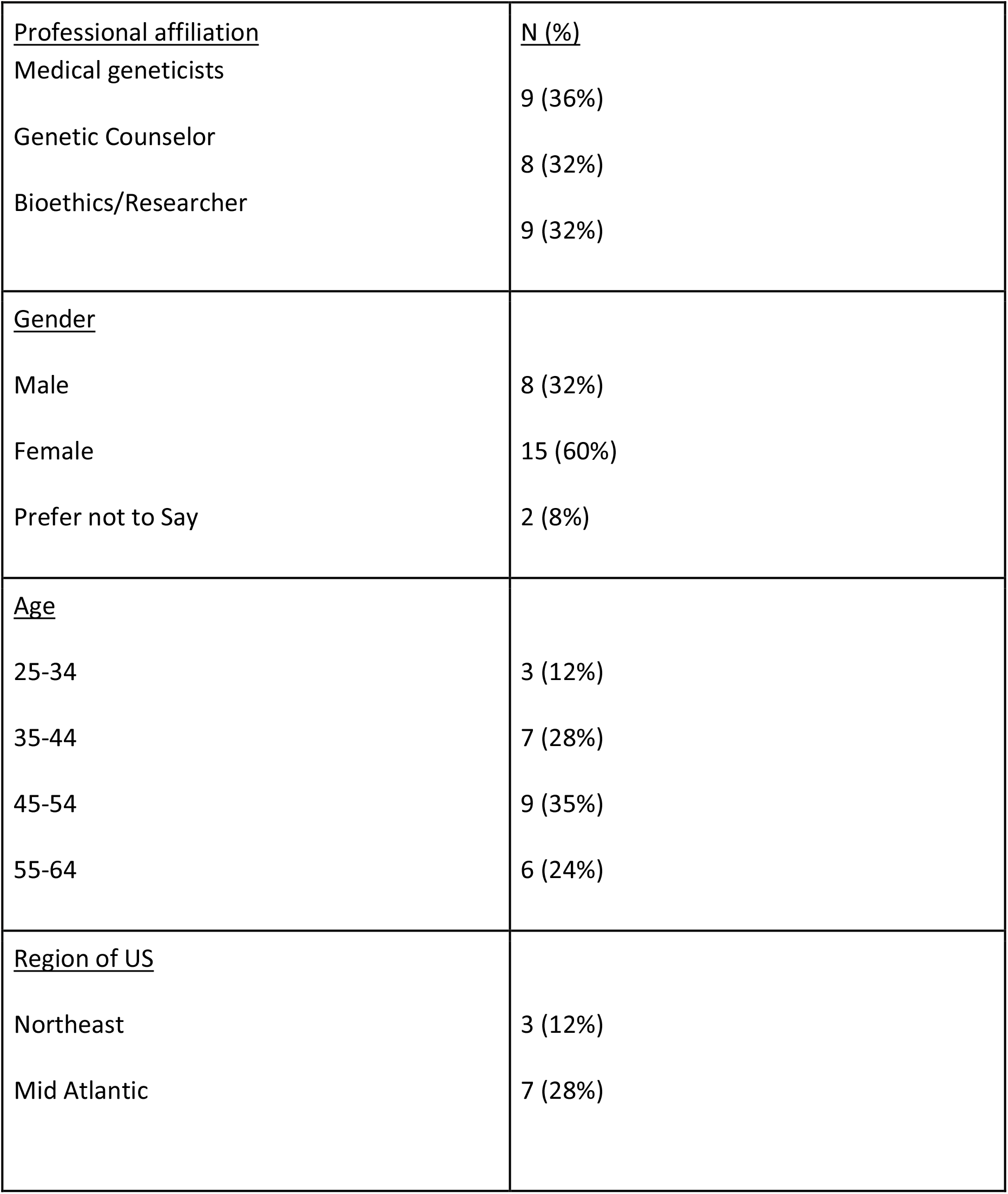
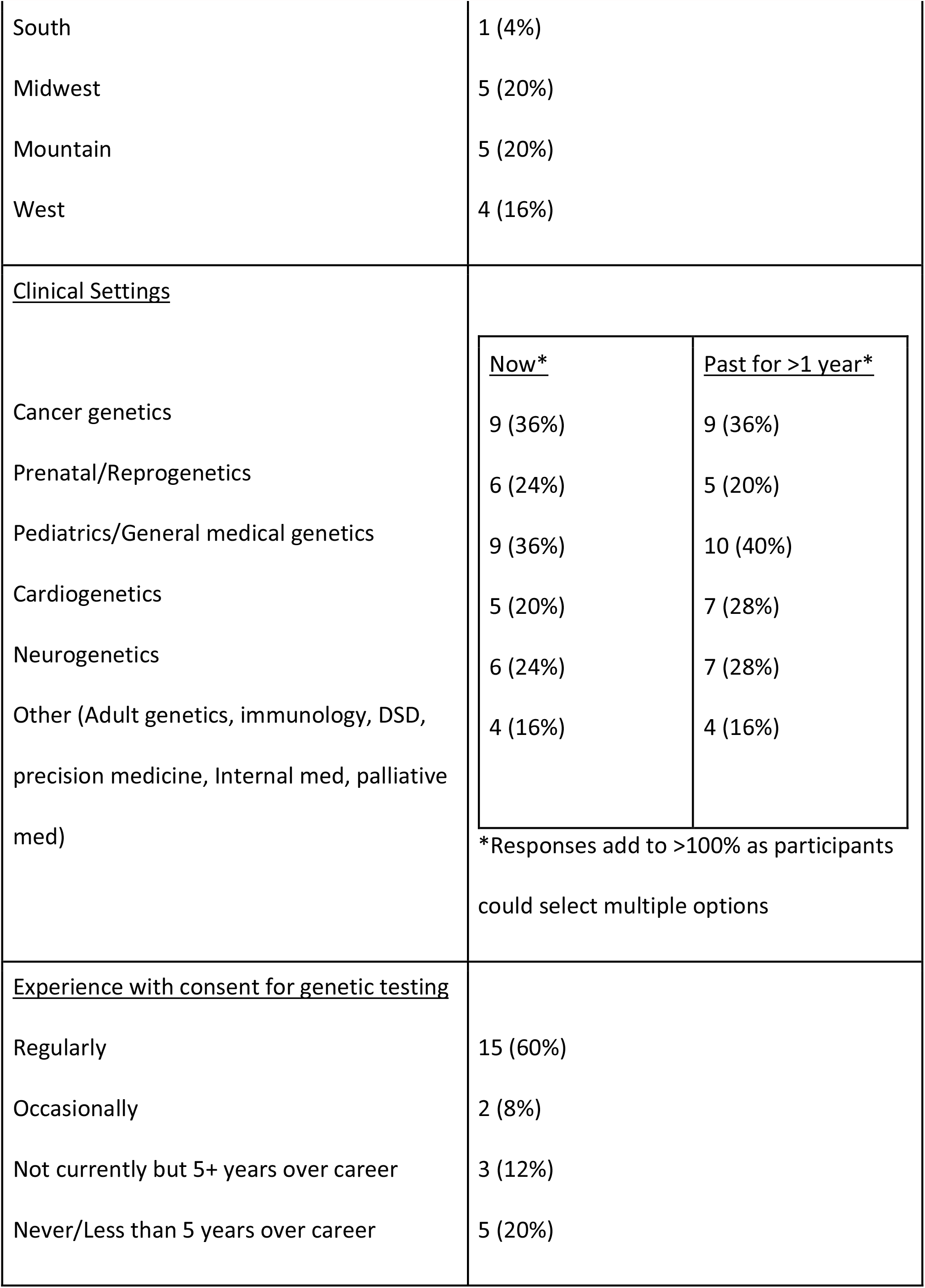
Demographics of Delphi Participants (N=25)

Table 2 shows the levels of agreement for the highest ranking concepts on each of the two surveys. Supplemental Table 1 shows the mean levels of agreement and rankings for each of the original 77 concepts plus the 12 additional concepts that were ranked on Survey 2.

**Table 2:** Most highly ranked concepts from Delphi Process.

| Concept | Survey 1<br>N=25<br>Mean ± SD | Survey 2<br>N=23<br>Mean ± SD | Comments |
| --- | --- | --- | --- |
| What is the condition we are testing for?<br>You may learn the cause of the indication we are testing for<br>Why are we doing the test | 4.76 ± .523<br>4.12 ± .881<br>Added in comments | 4.91 ± .288<br>4.30 ± .765<br>4.22 ± 2.07 | Combined in final list as “Why are we doing this test and what does it look for” |
| What results will be returned (generally)? | 4.68 ± .557 | 4.70 ± .559 | |
| How, if at all, will management be impacted by the results?<br>There may be an impact on your personal health through a diagnosis<br>We may have information on how to screen/treat some identifiable conditions | 4.40 ± .707<br>4.36 ± .810<br>4.16 ± .850 | 4.43 ± .590<br>4.50 ± .514<br>3.89 ± .583 | Combined in final list as “How, if at all, will your prognosis and management (including health screening) be impacted by the results?” |
| Genetic testing is always voluntary (optional) /<br>You have the right not to know about your genetic status | 4.20 ± .707<br>4.04 ± .978 | 4.22 ± .671<br>3.91 ± .996 | Combined in final list as “Genetic testing is always voluntary” |
| A diagnosis may also impact your family in different ways (their health, emotions or relationships)<br>You may learn unexpected information about family relationships<br>How to share with family | 4.12 ± .927<br>4.04 ± .790<br>Added in comments | 4.00 ± .686 (broadly),<br>3.96 ± .706<br>3.96 ± 1.107 | Combined in final list as “The results may also impact your family in different ways (their health, emotions, or relationships), and you may want to share the results.” |
| What other types of results will potentially be returned, and options for choice (such as secondary findings)? | 4.04 ± .790 | 4.17 ± .650 | |
| To whom the results will be reported? | 4.00 ± .816 | 4.00 ± .739 | |
| What are the limitations of the test?<br>Will there be further testing if no answer? | 3.84 ± .800<br>Added in comments | 3.96 ± .767<br>4.48 ± .805 | Represented in top 5 on survey 1; 52% (12/23) support adding in comments on S2 |
| There is a potential risk for genetic discrimination<br>There may be risks for discrimination or stigma (insurance, etc)<br>GINA and relevant state laws provide some protection. | 3.08 ± .997<br>3.91 ± .949<br>3.76 ± .926 | 3.52 ± .790<br>3.61 ± .778<br>3.87 ± .757 | All three items highly ranked in top 5 on S1. 43% (9/21) supported adding at least one of the items in comments on S2. Items combined and added to final list as "There is a potential risk for genetic discrimination and/or stigma/GINA and relevant state laws provide some protection" |

After presenting the list of highly ranked concepts in Survey 2, participants were asked if they felt there were any concepts that were missing (N=13 said “yes”) or could be removed (N=8 said “yes”). Twelve participants (52%) made open ended comments regarding items they felt were missing from the critical concept list. Concepts mentioned by more than one participant as missing from the list included: the uncertainty of benefit from genetic testing, general test limitations, how likely it would be for the test to find ‘an answer,’ how data is used and by whom, and issues around insurance and/or discrimination. Concepts mentioned as potentially removable included testing being voluntary, information about management, and the notion that some of the concepts on the list were overlapping.

Sixteen participants (70%) wrote comments regarding the list in general. Ten participants were supportive of the proposed list.

> *“I agree that it covers the concepts that most patients want to know about genetic testing*.*”*
>
> *“I think the list is helpful and focused. It should help improve the efficiency of obtaining consent and will probably improve the understanding of the people getting testing by simplifying the process*.*”*
>
> *“The list should allow for an informed decision. Some patients/families will have additional needs/interests which can be addressed during time allotted for questions*.*”*

Two added they had no further comments, and one highlighted that the survey and their responses “underlines the challenges in streamlining consent.” Two participants highlighted items that they still felt were missing from the proposed concept list: the manner in which the sample will be obtained (and its respective invasiveness) and how potential results will be interpreted.

Finally, participants were asked to review the list of 18 second tier concepts that were identified after the first survey. The most highly endorsed potential concepts were test limitations (12/23; 52%), variant interpretation will change over time (10/23, 44%), and the potential for genetic discrimination or stigma (9/23, 39%). Of note, six people, at least some of whom overlapped with those who endorsed including “genetic discrimination or stigma” also endorsed “GINA [Genetic Information Nondiscrimination Act] and relevant state laws provide some protection”. Most of the comments regarding GINA/discrimination issues stressed that there are important issues that people may not know about, and may actively wonder or worry about, that should be discussed prior to testing.

> *“I think this [GINA] can be wrapped into ‘potential risks for genetic discrimination and stigma, but patients should understand what is/isn’t protected legally before proceeding with testing*.*”*

### Development of the final consensus list

Based on two rounds of survey results, we developed a list of core concepts to evaluate and discuss within the CADRe workgroup (Table 2; Figure 1). Eight concepts met our criteria for being highly rated on both surveys (mean scores >4.0 and being most highly ranked among the top 5 within each group). One concept met the criteria for mean scores >4.0 on both surveys, but was not among the most highly ranked concepts (“You may learn the cause of the indication for which testing was done”). All of these items demonstrated < 0.18 change in mean score between the two surveys, demonstrating consistency in views among study participants. Two additional concepts that were added in response to open ended comments on the first survey had a mean score >4.0 on Survey 2: “Why are we doing the test” (mean 4.22) and “Will there be further testing if no answer [on the genetic test]” (mean 4.48). Two additional concepts that had been highly ranked in the top 5 on Survey 1 increased their ratings to >4.0 on Survey 2 such that they met both core criteria for being highly ranked: “We may have information on how to screen/treat some identifiable conditions” (3.89 to 4.16) and “You may learn unexpected information about family relationships” (3.96 to 4.04). One additional concept from the second tier concepts “What are the limitations of the test” demonstrated a slight increase in mean score (from 3.84 to 3.95) but demonstrated 52% support for inclusion (n=12/23) when respondents were specifically queried, and the example scenarios often had unsolicited comments that this topic was missing, so it was retained for inclusion on the final concept list.

**Figure 1:**
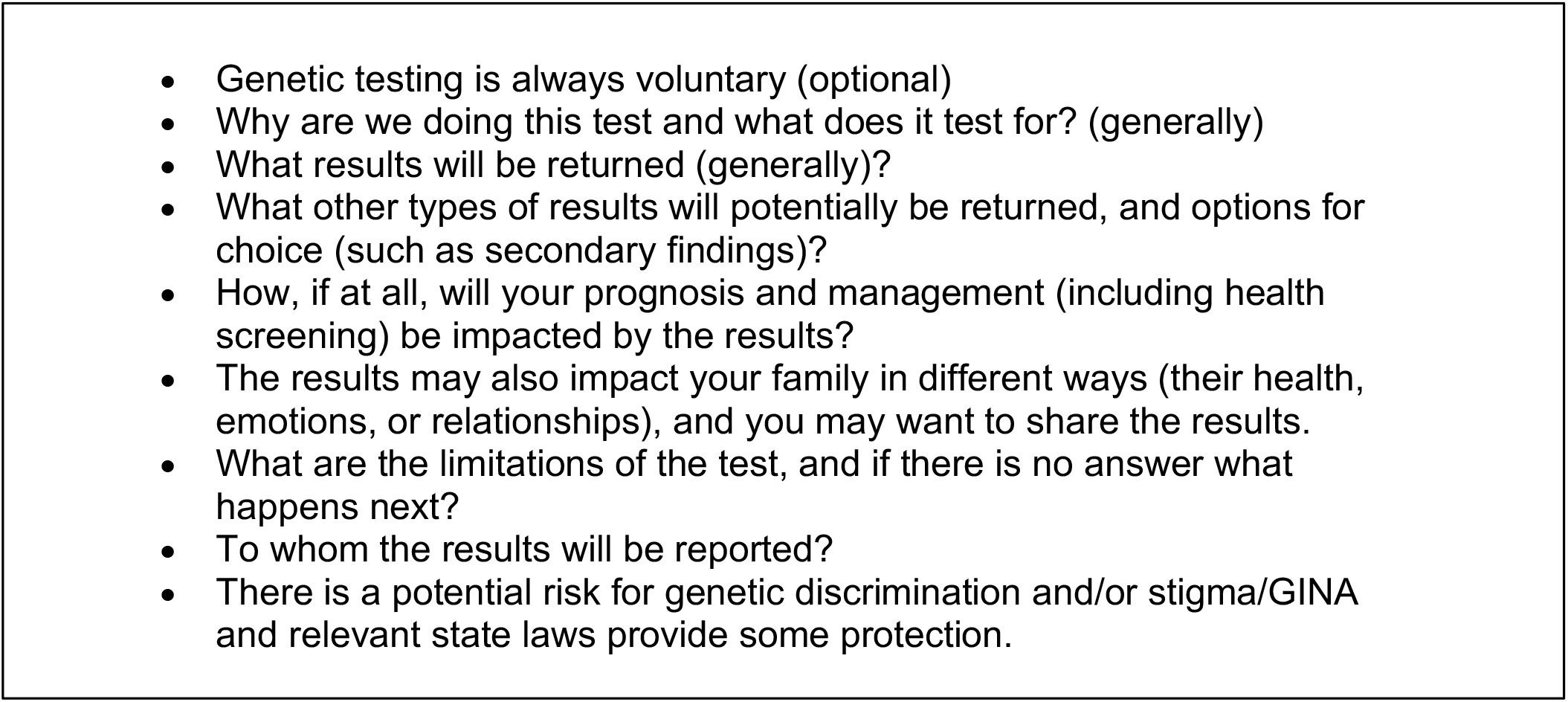
Final list of ‘necessary and critical’ concepts for informed consent for genetic testing.

## DISCUSSION

Over the past several years, the CADRe workgroup has taken up the question of how to “operationalize new approaches to provide patients with the education and support that each needs and desires when undergoing genetic testing.” ^23^. Prior studies of patients suggest that they desire a more straightforward and understandable consent process ^16,23,26^ with more specific and tailored information provided when test results are disclosed. However, while much discussion and debate has occurred about the concepts to include as part of pre-test informed consent around genetic testing, we are not aware of previous studies that operationalize various approaches to informed consent in this concrete way. Our current study developed consensus on a small list of content areas that experts with significant experience obtaining informed consent for genetic testing (geneticists, genetic counselors, and bioethicists who conduct research on informed consent for genetic testing) deemed “necessary and critical” to the process. Having done so, our results suggest that it is feasible to develop a narrow list of concepts that is not tied to any specific condition and that could serve as a ‘starting place’ for clinicians to discuss how best to facilitate informed consent discussions before genetic testing.

This study highlighted the following concepts as critical (Figure 1): the voluntariness of testing, the general reasons why the test is being performed and what is being tested for; the types of results returned; the limitations of testing; to whom the results will be reported; the impact results may have on prognosis, management, and family dynamics; and potential for discrimination and stigmas. While focused on clinical genetic testing, many of these mirror concepts required as part of a research consent: voluntariness of testing, the nature and purpose of the test, and the potential benefits and risks (including in this case the impact on self and family, and the potential for discrimination and stigma). This is also highly consistent with our own prior studies that have shown patients and families preferred to understand what is being tested for and why, what management changes might occur depending on test results, and the insurance implications. Similarly, non-genetics providers also prioritized reasons for testing and the potential impact on management, as well as test limitations and the likelihood of a positive result ^23^.

We propose that this list is tractable for both patients and clinicians, and provides all clinicians, whether genetics specialists or not, a focal point to begin the discussion about genetic testing with a patient. This concept list does not imply that only these concepts are important, but rather prioritizes a ‘most critical’ list as a starting place for discussion, and to avoid overwhelming patients with information that is potentially superfluous to their decision making process. It also recognizes that the full genetic counseling process, which often includes information regarding the logistics of sample collection and return of results, goes beyond the concepts that may be required to allow patients to make informed decisions about genetic testing. Given that our prior work suggests that for most predictive genetic testing an abbreviated (‘targeted discussion’) approach would be feasible and appropriate for many testing indications ^24^, this list would provide guidance to clinicians regarding what concepts might be included in a pre-test discussion about genetic testing. Additionally, having a broad framework of consent concepts that is agnostic to the genetic condition or gene for which testing is provided would potentially allow a clinician of any specialty background to feel prepared to identify the patient specific issues, including clinical testing indication and personal and family history that would allow tailoring the content and triaging referrals appropriately.

We anticipate there will be some genetics providers who feel uncomfortable with any concept list that attempts to prioritize concepts for informed consent. The recent US-based 21st Century Cures Act Information Blocking Rule (https://www.federalregister.gov/documents/2020/05/01/2020-07419/21st-century-cures-act-interoperability-information-blocking-and-the-onc-health-it-certification), along with laws that allow for direct release of genetic testing results to patients, may heighten this concern, making some providers worry that it is even more critical to provide as much information as possible during the pre-test consent conversation. Rather, we would emphasize that tailoring a pre-test genetic testing discussion to these points allows a patient either to make an informed decision or to request further information, and that information beyond these points may be best suited to a post-test conversation when providers can contextualize the results and their implications in a focused manner. By shortening the pre-test communication process, we expect that patients will still be able to make informed decisions about genetic testing, providers will feel more comfortable that they are covering the key concepts, and access may be improved through both the broadening pool of providers who are able to facilitate the genetic testing process as well as the shorter time frame required, allowing more patients to be seen in relevant clinics.

### Limitations

As is the case in most Delphi studies, the results of this study provide insight into what ‘experts’ think about a particular topic. Our study characterized experts as US based genetics care providers (geneticists and genetic counselors) with more than 5 years of experience obtaining consent for genetic testing, as well as bioethics researchers who research and write about the topic. While our demographics suggest we obtained a range of opinions from people who practice in different clinical areas, different geographic parts of the USA, and with different amounts of experience, it is possible that the views of these experts do not represent the broader professional attitudes, or even the attitudes that patients and families would have towards the same concepts.

## Conclusions

Our study has shown that it is possible to reach expert consensus on a minimal list of concepts that are critical to facilitate informed consent in patients considering genetic testing, and we anticipate the medical genetics community will robustly debate the concepts. Our next steps include determining if a broader sample of medical geneticists and genetic counselors agree that these concepts would allow patients to make informed decisions in a variety of clinical scenarios and indications (Borensztein et al., 2020) and assessing whether these views of critical concepts are supported by patients and families who have previously undergone genetic testing for conditions including familial hypercholesterolemia (FH), which CADRe has classified as a brief communication approach to consent ^24^, and for adult onset neurodegenerative conditions, which we have proposed occur through either traditional genetic counseling or targeted discussions depending on the testing indication ^24^.

## Data Availability

The entire set of summarized data from both surveys is available in the supplemental table.

## DATA AVAILABILITY

The entire set of summarized data from both surveys is available in the supplemental table.

## ACKNOWLEDGEMENTS

ClinGen is primarily funded by the National Human Genome Research Institute (NHGRI), through the following three grants: U41HG006834, U41HG009649, U41HG009650. ClinGen also receives support for content curation from the Eunice Kennedy Shriver National Institute of Child Health and Human Development (NICHD), through the following three grants: U24HD093483, U24HD093486, U24HD093487. The content is solely the responsibility of the authors and does not necessarily represent the official views of the National Institutes of Health

Thanks to Erin Ramos and Nicole Lockhart at NHGRI, and all the remaining members of the CADRe workgroup: Laura Hercher, Howard P. Levy, Julianne M. O’Daniel, Juliann M. Savatt, Melissa Stosic, Hannah Wand. The work described was based on input from CADRe working group, with additional members having contributed conceptually at various stages in the development of the rubrics, including: Kyle B. Brothers, Louanne Hudgins, Seema M. Jamal, Dave Kaufman, and Myra I. Roche. Finally, we are grateful to members of the Stanford Center for Biomedical Ethics writing seminar for comments on an earlier version of this paper.

## AUTHOR INFORMATION

Conceptualization: K.E.O, M.L.G.H, A.H.B, W.A.F, C.R.C; Data Curation: K.E.O, M.B.; Formal Analysis: K.E.O.; Funding Acquisition: K.E.O, M.L.G.H, A.H.B, W.A.F; Investigation: K.E.O, M.B.; Methodology: K.E.O, M.L.G.H, H.L.P, A.H.B, M.E.S, W.R.U, K.E.W., C.R.C; Project administration: K.E.O, M.B., E.P.T; Supervision: K.E.O, M.L.G.H, A.H.B; Visualization: K.E.O, M.B., E.P.T; Writing - original draft: K.E.O, M.B.; Writing - review and editing: K.E.O, M.B., M.L.G.H, A.H.B, W.A.F, H.L.P, M.E.S, E.P.T, W.R.U, K.E.W., C.R.C.

## ETHICS DECLARATION

The Stanford IRB reviewed this study as exempt under category 2, and all aspects adhered to the principles in the Declaration of Helsinki. All participants reviewed an informed consent document before choosing to participate in this study.

---

Supplemental table 1. Full results from Delphi Survey Means and Rankings (77 Original Concepts + 12 Added Concepts)

