## Supplemental Table 1 for "Defining the Critical Components of Informed Consent for Genetic Testing: A Delphi Study"

Supplemental Table 1. Full results from Delphi Survey Means and Rankings (77 Original Concepts + 12 Added Concepts)

| **Concept** | **Survey 2 (N=23) Group Mean ± SD** | **Survey 1 (N=25) Group Mean ± SD** | **Mode S1** | **Total ranked in top 5 on S1** | **Rank #1** | **Rank #2** | **Rank #3** | **Rank #4** | **Rank #5** |
|---|---|---|---|---|---|---|---|---|---|
| ***What is the condition we are testing for?*** | ***4.91 ± .288*** | ***4.76 ±.523*** | ***5*** | ***23*** | ***19*** | ***-*** | ***-*** | ***1*** | ***3*** |
| ***What results will be returned (generally)?*** | ***4.70 ± .559*** | ***4.68 ± .557*** | ***5*** | ***17*** | ***2*** | ***5*** | ***7*** | ***3*** | ***-*** |
| ***There may be an impact on your personal health through a diagnosis*** | **4.50 ± .514** | ***4.36 ± .810*** | ***5*** | ***23*** | ***13*** | ***4*** | ***3*** | ***1*** | ***2*** |
| ***How, if at all, will management be impacted by the results?*** | ***4.43 ± .590*** | ***4.40 ± .707*** | ***4*** | ***15*** | ***1*** | ***10*** | ***2*** | ***1*** | ***1*** |
| ***You may learn the cause of the indication for which testing was done*** | ***4.30 ± .765*** | ***4.12 ± .881*** | ***5*** | ***3*** | ***-*** | ***1*** | ***-*** | ***1*** | ***1*** |
| ***Genetic testing is always voluntary (optional)*** | ***4.22 ± .671*** | ***4.20 ± .707*** | ***4*** | ***12*** | ***5*** | ***3*** | ***1*** | ***2*** | ***1*** |
| ***What other types of results will potentially be returned, and options for choice (such as secondary findings)?*** | ***4.17 ± .650*** | ***4.04 ± .790*** | ***4*** | ***14*** | ***1*** | ***2*** | ***2*** | ***5*** | |

| | | | | | | | | | |
|---|---|---|---|---|---|---|---|---|---|
| ***A diagnosis may also impact your family (broadly)*** | **4.00 ± .686** | ***4.12 ± 9.27*** | ***5*** | ***15*** | ***1*** | ***5*** | ***4*** | ***4*** | ***1*** |
| ***To whom the results will be reported?*** | ***4.00 ± .739*** | ***4.00 ± .816*** | ***4*** | ***10*** | ***1*** | ***3*** | ***3*** | ***2*** | ***1*** |
| ***You may learn unexpected information about family relationships*** | ***3.96 ± .706*** | ***4.04 ± .790*** | ***4*** | ***23*** | ***8*** | ***1*** | ***4*** | ***6*** | ***4*** |
| What results does one have the choice to receive or decline? | 3.96 ± .665 | 3.96 ± .735 | 4 | 12 | 5 | 2 | 2 | 1 | 2 |
| What are the limitations of the test? | 3.96 ± .767 | 3.84 ± .800 | 4 | 10 | - | - | 1 | 2 | 7 |
| ***You have the right not to know about your genetic status*** | *3.91 ± .996* | ***4.04 ± .978*** | ***5*** | ***6*** | ***1*** | ***1*** | ***1*** | ***-*** | ***3*** |
| ***We may have information on how to screen/treat some identifiable conditions*** | 3.89 ± .583 | ***4.16 ± .850*** | ***4*** | ***14*** | ***1*** | ***8*** | ***2*** | ***2*** | ***1*** |
| For some conditions you may want to personally prepare or change things about your life (education or job plans, reproductive plans) | 3.89 ± .900 | 3.92 ± .862 | 4 | 10 | 2 | 1 | 4 | - | 3 |
| How is the condition treated or managed? | 3.87 ± .694 | 3.8 ± .913 | 4 | 2 | - | 2 | - | - | - |
| GINA and relevant state laws provide some protection | 3.87 ± .757 | 3.76 ± .926 | 4 | 19 | - | 10 | 3 | 2 | 4 |

| | | | | | | | | | |
|---|---|---|---|---|---|---|---|---|---|
| There may be more than one disease risk | 3.83 ± .834 | 3.76 ± .926 | 4 | 3 | - | - | 1 | - | 2 |
| What is the likelihood of discovering cause of primary indication | 3.78 ± .671 | 3.80 ± .707 | 4 | 5 | - | 1 | 2 | 2 | **-** |
| Family members may learn unexpected health information | 3.74 ± .689 | 3.92 ± .862 | 4 | 15 | 2 | 8 | 2 | - | 3 |
| How ‘accurate’ are these results? | 3.74 ± .915 | 3.84 ± .987 | 3 | 8 | 3 | 2 | 1 | 1 | 1 |
| Ways variants can be interpreted: pathogenic, benign, VUS | 3.74 ± .752 | 3.76 ± .879 | 3 | 7 | **-** | **-** | ***4*** | ***2*** | ***1*** |
| We don’t yet understand all variants and their impact on disease risk | 3.74 ± 1.096 | 3.72 ± 1.100 | 4 | 7 | - | - | 2 | 3 | 2 |
| Variant interpretation may change over time | 3.70 ± .974 | 3.96 ± .841 | 4 | 6 | - | 4 | 1 | 1 | - |
| There may be a chance of false positive or false negative result | 3.70 ± .974 | 3.80 ± 1.000 | 4 | 5 | 1 | 1 | 2 | 1 | - |
| Your family may benefit from knowing their personal risks | 3.67 ± .594 | 3.84 ± .850 | 4 | 11 | 1 | - | 1 | 7 | 2 |
| How will the results be reported (phone, in person)? | 3.65 ± .982 | 3.76 ± .831 | 4 | 9 | - | 2 | 1 | 5 | 1 |
| There may be risks for discrimination or stigma (insurance, etc) | 3.61 ± .778 | 3.91 ± .949 | 4 | 21 | 12 | 5 | 4 | - | - |
| What types of results will NOT be returned | 3.57 ± 1.08 | 3.60 ± 1.155 | 4 | 7 | - | - | 1 | 4 | 2 |

| | | | | | | | | | |
|---|---|---|---|---|---|---|---|---|---|
| Degree of risk from results and impact | 3.52 + .898 | 3.80 + .957 | 3 | 6 | 1 | 1 | 1 | 1 | 2 |
| How likely will the test be negative even if the condition is familial (we didn't find the real cause)? | 3.52 + .790 | 3.56 + .870 | 3 | 4 | - | - | 1 | 1 | 2 |
| How long will it take to receive the results? | 3.52 + .680 | 3.56 + .821 | 3 | 9 | 1 | 2 | 3 | 2 | 9 |
| Differentiating single gene/panel/exome testing (as relevant)? | 3.52 + .665 | 3.52 + .963 | 4 | 1 | - | - | 1 | - | - |
| How do you get the sample (e.g. blood, buccal)? | 3.52 + 1.039 | 3.48 + 1.122 | 3 | 10 | 5 | 2 | 1 | 1 | 1 |
| Other tests (more focused or more broad reaching) may (or may not) be available | 3.52 + .790 | 3.48 + .872 | 3 | 1 | - | - | - | - | 1 |
| There is a potential risk for genetic discrimination | 3.52 + .790 | 3.08 + .997 | 2 | 21 | 8 | 2 | 6 | 3 | 2 |
| *You may feel that having a specific genetic diagnosis is personally valuable information | 3.50 + 1.043 | 3.64 + .907 | 4 | 13 | 3 | 4 | 3 | - | 3 |
| Your family may benefit from knowing their reproductive risks | 3.50 + .618 | 3.64 + .810 | 4 | 8 | - | 1 | - | - | 7 |
| What is the likelihood of other findings (incidental/secondary ) | 3.48 + .846 | 3.56 + .821 | 3 | 2 | - | - | 1 | - | 1 |

| | | | | | | | | | |
|---|---|---|---|---|---|---|---|---|---|
| How will reinterpreted results be returned to the patient/client? | 3.48 ± .947 | 3.56 ± 1.121 | 4 | 1 | - | - | 1 | - | - |
| Sometimes we will not be certain about the potential benefits | 3.48 ± .982 | 3.44 ± 1.083 | 3 | 0 | - | - | - | - | - |
| Your family may have positive health impacts from this test | 3.44± .616 | 3.72 ± .792 | 4 | 7 | 1 | - | 1 | 3 | 2 |
| It could be hard to learn about unexpected or untreatable conditions or those with unexpected or unclear prognosis | 3.44 ± .705 | 3.57 ± .896 | 4 | 16 | 2 | 9 | 5 | - | - |
| Results may vary in how immediately they matter | 3.44 ± .922 | 3.40 ± .816 | 3 | 7 | 2 | 2 | 1 | - | 2 |
| Is any individual data entered into public databases | 3.43 ± .945 | 3.40 ± 1.041 | 4 | 9 | 1 | 3 | - | 3 | 2 |
| The test may identify multiple conditions | 3.39 ± .502 | 3.68 ± .900 | 4 | 12 | 1 | - | 5 | 5 | 1 |
| Will family members' results be reported (if used)? | 3.30 ± .822 | 3.52 ± .918 | 3 | 2 | - | - | 1 | 1 | - |
| How will the patient/client know if results have been reinterpreted? | 3.30 ± .876 | 3.48 ± 1.122 | 4 | 1 | - | - | - | - | 1 |

| | | | | | | | | | |
|---|---|---|---|---|---|---|---|---|---|
| What is the mode(s) of inheritance of condition? | 3.30 ± .635 | 3.40 ± .816 | 3 | 3 | - | 1 | 2 | - | - |
| Your genetic data could identify you | 3.28 ±.669 | 3.57 ± .843 | 3 | 15 | 3 | 4 | 8 | - | - |
| There may be different implications if you are employed by business with fewer than 15 people, US Military, or the Federal Government | 3.22 ± .850 | 3.40 ±1.000 | 3 | 8 | - | - | 5 | 1 | 2 |
| Where will the results be placed or stored (e.g. EMR, patient portal)? | 3.22 ± .902 | 3.36 ± .952 | 3 | 4 | - | 1 | 3 | - | - |
| You may have challenging emotional responses such as anxiety, distress, surprise, confusion | 3.17 ±.857 | 3.61 ±.783 | 3 | 13 | 3 | 5 | 5 | - | - |
| Samples from other family members may help interpret the test more accurately | 3.13 ±.757 | 3.44 ± .917 | 3 | 7 | 1 | 2 | 1 | 1 | 2 |
| Under what circumstances is a reanalysis initiated and by whom? | 3.13 ± .968 | 3.28 ±1.021 | 3 | 5 | 1 | - | - | 2 | 2 |
| Some people may feel more motivated to change their health behaviors after genetic testing results | 3.11 ±.758 | 3.16 ± .850 | 3 | 5 | - | - | 1 | 3 | 1 |

| | | | | | | | | | |
|---|---|---|---|---|---|---|---|---|---|
| Effects of variants can vary between family members and between families | 3.09 ± .996 | 3.40 ± 1.000 | 3 | 1 | - | - | - | - | 1 |
| What is the likelihood of variants we don't understand (VUS) | 3.09 ± .793 | 3.24 ± 1.091 | 2 | 3 | - | - | - | 2 | 1 |
| How is data sharing decided and by whom? | 3.09 ± .949 | 3.12 ± 1.092 | 3 | 12 | 2 | 1 | 1 | 4 | 4 |
| Variants may increase or decrease the risks | 3.04 ± .878 | 3.40 ± 1.118 | 4 | 0 | - | - | - | - | - |
| Variants are complicated to interpret | 3.00 ± .674 | 3.48 ± .823 | 3 | 0 | - | - | - | - | - |
| Where and how long will the data reports be kept? | 2.96 ± .825 | 2.80 ± .816 | 3 | 5 | 2 | - | 2 | - | 1 |
| What gene(s) are included in the test? | 2.96 ± .976 | 2.76 ± .879 | 2 | 2 | - | 2 | - | - | |
| What is the likelihood of other uncertainties in results? | 2.91 ± .733 | 3.00 ± .957 | 3 | 0 | - | - | - | - | - |
| What will happen to the results if patient dies? | 2.70 ± .926 | 3.04 ± 1.020 | 3 | 0 | - | - | - | - | - |
| What is DNA/Genes/Chromosomes (as relevant)? | 2.65 ± .714 | 2.92 ± .997 | 2 | 1 | 1 | - | - | - | - |
| How often will a reanalysis occur? | 2.65 ± .647 | 2.96 ± .978 | 3 | 0 | - | - | - | - | - |
| Are there any physical risks in obtaining the sample? | 2.65 ± .832 | 2.96 ± 1.136 | 2 | 0 | - | - | - | - | - |

| | | | | | | | | | |
|---|---|---|---|---|---|---|---|---|---|
| Who can request data or reports if the patient dies? | 2.65 ± .935 | 2.88 ± .971 | 2 | 6 | - | - | 1 | 4 | 1 |
| What else will the samples be used for and by whom? | 2.61 ± .783 | 3.00 ± 1.258 | 2 | 2 | - | - | - | 1 | 1 |
| Who can request 'raw data', and how? | 2.57 ± .945 | 2.76 ± .970 | 2 | 3 | 1 | - | - | 1 | 1 |
| "Everyone has some mutations"? | 2.43 ± 0.896 | 2.56 ± .917 | 2 | 0 | - | - | - | - | - |
| Is any aggregate data entered into public databases? | 2.39 ± .839 | 2.44 ± .768 | 2 | 0 | - | - | - | - | - |
| How, where and how long will the sample be kept? | 2.35 ± .647 | 2.40 ± .764 | 2 | 2 | 1 | - | - | 1 | - |
| There may be physical risks to getting the sample* | 2.33 ± 1.085 | 2.26 ± 1.096 | 2 | 4 | 3 | - | 1 | - | - |
| What is the name of the lab that will run the sample? | 2.17 ± .778 | 2.16 ± .850 | 2 | 2 | - | - | - | - | 5 |
| Will the sample ever be used for quality control? | 1.87 ± .757 | 1.92 ± .759 | 2 | 0 | - | - | - | - | - |
| Technical data on how the sample is run | 1.74 ± .619 | 1.68 ± .557 | 2 | 0 | - | - | - | - | - |
| ****Q's only on S2 listed below** | | | | | | | | | |
| **Will there be further testing if this test doesn't give answer?** | **4.48 ± .805** | | **4** | | | | | | |

| | | | |
|---|---|---|---|
| **Why are we doing this test?** | **4.22 ± 2.066** | | **5** |
| How to share results with family members who may be at risk | 3.96 ± 1.107 | | 5 |
| You may be disappointed if this test does not yield an answer for why you are sick | 3.78 ± 1.085 | | 3 |
| What data might be shared? | 3.74 ± 1.214 | | 4 |
| What about negative impact on family | 3.48 ± 1.534 | | 3 |
| The differences between raw data, interpreted data and interpretive summaries | 3.43 ± .896 | | 4 |
| Your personal identity may be compromised (e.g. identity theft, criminal) | 3.30 ± 1.363 | | 3 |
| There is potential for disagreement about what to do with a genetic diagnosis in a family. This can lead to interpersonal conflict | 3.26 ± 1.096 | | 3 |
| How data (not just sample) will be shared or stored, particularly from a commercial | 3.22 ± 1.242 | | 3 |

| | | | |
|---|---|---|---|
| perspective (sales, transfers) | | | |
| Group privacy considerations | 3.22 ± 1.166 | | 3 |
| How to access raw data and its interpretations | 2.83 ± 1.029 | | 3 |

Items are presented in rank order from highest to lowest means on S2. Items in bold demonstrated at least one round with mean scores ≥4.00

**Additional 12 concepts that were added to S2 after free-text comments on S1**

*Due to a technical glitch there are some questions that were only answered by N=18 (of 23) respondents in Survey 2.
